# Characterization and Prediction of Clinical Pathways of Vulnerability to Psychosis through Graph Signal Processing

**DOI:** 10.1101/2020.06.11.20128769

**Authors:** Corrado Sandini, Daniela Zöller, Maude Schneider, Anjali Tarun, Marco Armando, Barnaby Nelson, Sumudu Rasangi Mallawaarachchi, G. Paul Amminger, John Farhall, Luke K. Bolt, Hok Pan Yuen, Connie Markulev, Miriam R. Schäfer, Nilufar Mossaheb, Monika Schlögelhofer, Stefan Smesny, Ian B. Hickie, Gregor Emanuel Berger, Eric Y.H. Chen, Lieuwe de Haan, Dorien H. Nieman, Merete Nordentoft, Anita Riecher-Rössler, Swapna Verma, Andrew Thompson, Alison Ruth Yung, Kelly A. Allott, Patrick D. McGorry, Dimitri Van De Ville, Stephan Eliez

## Abstract

There is a growing recognition that psychiatric symptoms have the potential to causally interact with one another. Particularly in the earliest stages of psychopathology dynamic interactions between symptoms could contribute heterogeneous and cross-diagnostic clinical evolutions. Current clinical approaches attempt to merge clinical manifestations that co-occur across subjects and could therefore significantly hinder our understanding of clinical pathways connecting individual symptoms. Network approaches have the potential to shed light on the complex dynamics of early psychopathology. In the present manuscript we attempt to address 2 main limitations that have in our opinion hindered the application of network approaches in the clinical setting. The first limitation is that network analyses have mostly been applied to cross-sectional data, yielding results that often lack the intuitive interpretability of simpler categorical or dimensional approaches. Here we propose an approach based on multi-layer network analysis that offers an intuitive low-dimensional characterization of longitudinal pathways involved in the evolution of psychopathology, while conserving high-dimensional information on the role of specific symptoms. The second limitation is that network analyses typically characterize symptom connectivity at the level of a population, whereas clinical practice deals with symptom severity at the level of the individual. Here we propose an approach based on graph signal processing that exploits knowledge of network interactions between symptoms to predict longitudinal clinical evolution at the level of the individual. We test our approaches in two independent samples of individuals with genetic and clinical vulnerability for developing psychosis.

## Introduction

Psychiatric disorders are remarkably complex. By the time an individual manifests a sufficient decline in quality of life to warrant consultation with a mental health professional he will often present a heterogeneous collection of multiple signs and symptoms. The urgent need to provide optimal clinical care, that is evidence-based and consistent across clinicians requires a systematic approach to address such complexity and heterogeneity [1, 2]. In particular, clinical practice involves massively reducing dimensionality of information, from the quantitation of up to hundreds of symptoms, to a much more limited number of potential treatment options. Current approaches to tackle complex clinical patterns in psychiatry have invariably merged together clinical manifestations that tend to co-occur across subjects. The inherent guiding principle is that if two symptoms co-occur in a sufficiently high proportion of patients, their clinical distinction becomes redundant for guiding clinical decision making. The prototypical example of this approach consists in establishing boundaries within which co-occurrence of psychiatric symptoms is sufficiently high to warrant a single diagnostic label [3]. This intuitive approach has proven extremely useful in increasing communicability and agreement across clinicians [1]. There is, however, growing concern that such categorical distinctions may be a step too far in reducing the complexity of mental health disturbances [4]. Indeed, diagnostic algorithms have demonstrated limited utility in guiding therapeutic decisions, which strongly supports the need for reform [5]. An alternative approach consists in progressively merging manifestations of mental health disturbances over progressively higher levels of complexity, on the basis of their empirically observed pattern of co-occurrence, providing a potentially more accurate representation of mental health phenomena [6, 7]. It remains however very much debated whether dimensional approaches would provide benefits in guiding clinical decision making [8]. Indeed while “dimension fit the data” it is still unclear whether “clinicians can fit dimensions” [8].

It has been argued that part of the dis-satisfaction towards both dimensional and categorical approaches to psychopathology may stem from the underlying conceptualization regarding the origins and nature of mental health disturbances [9, 10]. Indeed, the implicit assumption justifying the merging of psychiatric symptoms into sub-scales, dimensions or diagnoses, is that co-occurrence of symptoms is accounted for by existence of single underlying causal factors [10]. In psychiatry however, it is increasingly recognized that symptoms are not only passive expression of common underlying disease processes, but can in some cases represent active agents that have the ability to provoke their reciprocal emergence, through dynamic causal interactions [9, 10]. For instance, the observation that in patients with chronic psychosis, thought disorders tend to co-occur with social retreat could be explained by the fact that early sub-clinical paranoid ideation hindered the subsequent maintenance of functional social interactions, consistent with the concept of secondary negative symptoms [11]. Similarly, a causal association between early insomnia and subsequent mood disturbances could partially account for their co-occurrence in depressed patients [12]. Moreover, the emerging study of at-risk populations has increasingly demonstrated that the earliest manifestations of psychopathology are largely not specific to their corresponding clinical outcome [2, 13-15]. For instance, sub-threshold psychotic symptoms increase the likelihood of developing not only a full-blown psychotic disorder, but also mood, anxiety and substance use disorders [16, 17]. On the opposite, the presence of affective and amotivation symptoms strongly increase the likelihood of conversion to psychosis in individuals with psychotic symptoms [14, 18]. In the field of developmental psychopathology, cross-disorder interactions are probably particularly prominent, where they have been described as sequential comorbidity [19] [20]. From the perspective of pathophysiology, the extent of such cross-disorder interactions is largely incompatible with the idea that symptoms of a particular disorders emerge as a consequence of a series of separate and discrete causal factors. They rather suggest that different psychiatric symptoms can undergo dynamic interactions over time which could in turn account for emergence of the complex and heterogeneous clinical patterns, that are typically observed in individuals with mental health disturbances [13, 15]. From the clinical perspective, understanding and modeling pathways of interaction between symptoms carries tremendous potential, in terms of establishing prognosis and planning treatment strategies.

Network science is a rapidly expanding branch of mathematics dedicated to the study of graphs, which can be broadly defined as structures composed of discrete nodes that are connected by edges [21]. Applications of network science range from the study of networks of social interactions [22] to that of networks of biological interaction between genetic transcripts [23]. Moreover, network techniques are increasingly employed to study the interactions between symptoms of psychopathology [24, 25]. The most widely implemented paradigm has consisted of measuring correlations between different pairs of psychiatric symptoms in cross-sectional samples, reconstructing a network of symptoms-symptoms interactions[10, 24, 25]. Techniques of network science then allow to identify groups of symptoms, defined as clusters or modules, that are more densely interconnected with one another than with the rest of the symptoms in the network. Moreover, it is possible to identify symptoms that are disproportionately associated with the severity of other clinical manifestations, and that are said to have high network centrality. High centrality is commonly considered to reflect of a prominent causal role in influencing other symptoms. Network approaches are rapidly gaining popularity by demonstrating that exploiting high-dimensional granularity of clinical assessments can generate insights that would be missed if symptoms were merged in diagnoses or dimensions. Still, despite considerable promise, network approaches have to date largely remained confined to the laboratory. Below, we suggest that recent methodological advances made in the fields of dynamic network analysis and graph-signal-processing can help address 3 main obstacles to the clinical translation of network approaches to psychopathology.

The first shortcoming is that computational challenges have largely limited the application of network techniques to cross-sectional data. As a consequence, psychiatric symptoms networks typically lack the essential dimension of time. For instance, high centrality in a cross-sectional sample could imply that a symptom has an active role in broadly influencing subsequent clinical manifestations. However, an opposite but equally likely interpretation is that high centrality reflects the tendency of symptoms to be passively influenced by different prior psychiatric manifestations. To address this limitation we propose a Temporal Multilayer Symptom Network (TMSN) approach mutated from dynamic network analysis [26]. A TMSN, applied to developmental psychopathology, would consist of a first temporal layer composed of cross-sectional correlations between symptoms at a first baseline assessment. The subsequent network layers are composed of correlations between symptoms measured at longitudinal follow-ups. Such cross-sectional layers would be connected by longitudinal edges reflecting the association of symptom across time, namely which symptoms at baseline predicted which symptoms at follow-up. Analytic tools of network science could then allow the dissection of longitudinal disease pathways connecting manifestations of psychopathology over time [26]. For instance, it would be possible to dissect symptoms at baseline that broadly affect clinical manifestations at follow-up, and can be conceptualized as *gateways* of psychopathology, from symptoms at follow-up that are broadly affected by psychopathology at baseline, acting as *funnels* of psychopathology (See Figure 1).

**Figure 1.**
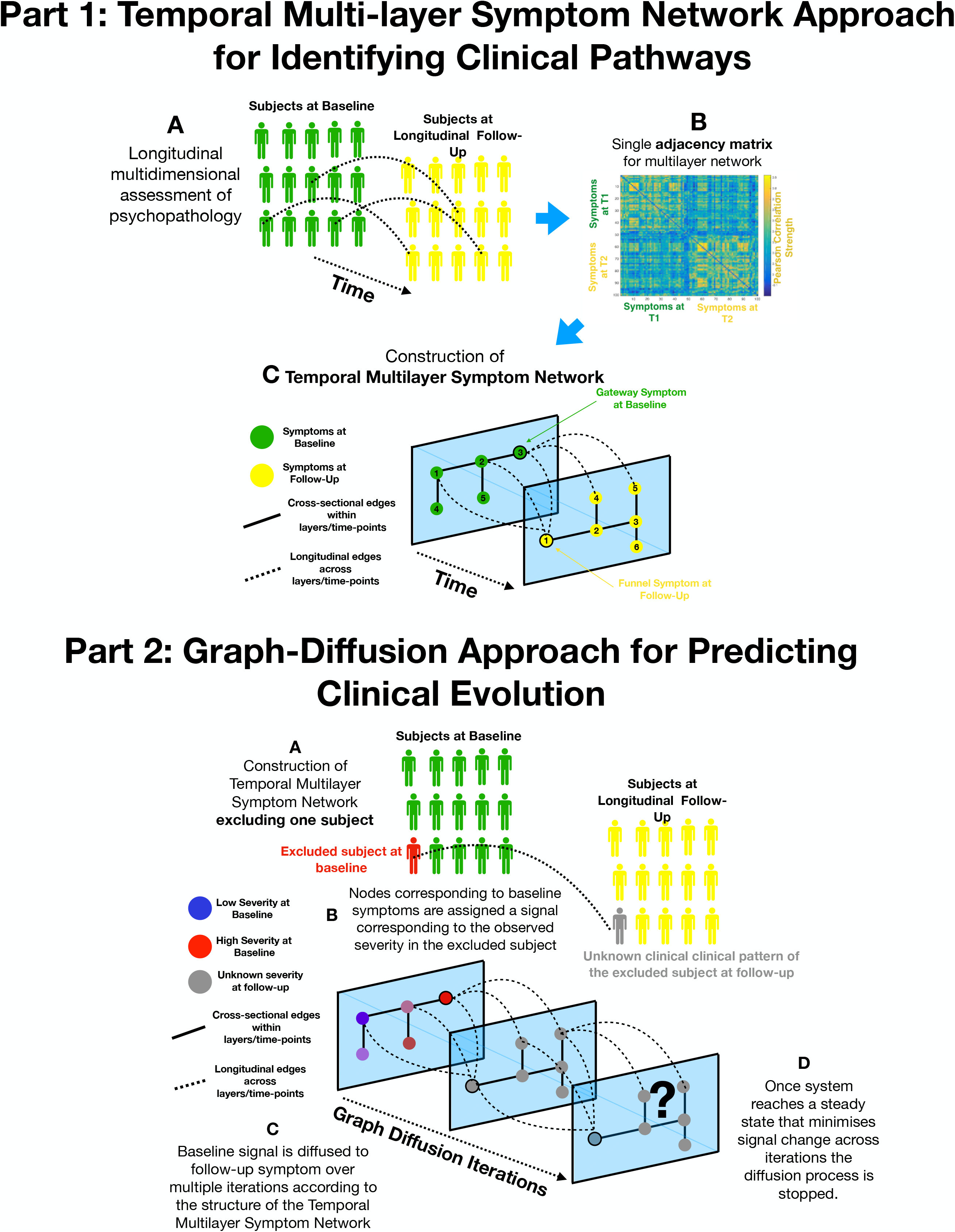
**Part 1:** Methodological pipeline for construction of Temporal Multi-Layer Symptom Network. **A:** Clinical assessment of multiple symptoms is performed for a cohort of participants over two time-points (Baseline and Follow-Up). **B:** A single adjacency matrix is constructed, containing cross-sectional correlations between symptoms measured at baseline and follow-up which are located respectively in the upper-left and lower right quadrants. The off-diagonal quadrant is composed of longitudinal correlations between each symptom at baseline and each symptom at follow-up. **C:** Graphical representation of the multilayer symptom network. A first network layer is composed of correlation between baseline symptoms, represented in green and a second layer is composed of correlations between symptoms at follow-up represented in yellow. The two cross-sectional layers are connected by longitudinal edges composed of correlations between baseline and follow-up symptoms represented as dashed lines. Such intuitive graphical representation is achieved empirically through topological embedding of symptoms according to dimensions derived from Eigen-decomposition of the multilayer adjacency matrix. Graph-theory is employed to identify longitudinal clinical pathways (shortest paths) connecting symptoms across temporal layers. Baseline symptoms that broadly influence symptoms at follow-up, have high longitudinal centrality, and can be conceptualized as gateways of psychopathology (schematically represented as symptom 3 at baseline). Follow-up symptoms with high longitudinal centrality are broadly influenced by symptoms at baseline, and can be conceptualized by funnels of psychopathology (schematically represented as symptom 1 at follow-up). **Part 2:** Methodological pipeline for Graph-Diffusion based prediction of clinical evolution. **A:** A Temporal Multilayer Symptom Network is reconstructed excluding the subject for whom clinical prediction is performed, in a leave-one-out cross-validation loop. **B:** In a graph-signal-processing framework node corresponding to baseline symptoms are assigned a signal that corresponds to their empirically observed severity in the excluded subject. At the beginning of the diffusion process severity of symptoms at follow-up is considered to be unknown and corresponding nodes are assigned a value of 0. **C:** Using a finite difference graph diffusion approach signal corresponding to the observed clinical pattern at baseline is diffused on the Temporal Multilayer Symptom Network. Compared to simple regression analysis prediction keeps into account both the structure of longitudinal correlations connecting layers of baseline and follow-up symptoms, which are represented schematically as dashed lines, and the structure of cross-sectional correlations between symptoms at follow-up. This diffusion approach leads to a progressive evolution of the predicted symptom pattern at follow-up over multiple diffusion iterations. The symptom pattern at baseline is considered to be known, and is hence re-initialized at each diffusion iteration. This is conceptually similar to modeling the spread of information in a social network as a function of friendship ties between individuals or the diffusion of temperature as a function of distance between spatial locations. **D:** For temperature, information or psychopathology, the diffusion algorithm will evolve the predicted signal until the system converges towards an equilibrium that minimizes signal change across time, at which point prediction is considered to be stable the iterative diffusion will be stopped. This process was repeated to predict symptom severity at follow-up for each subject included in the cohort, in a leave-one-our cross-validation loop.

A second limitation is that current representations of network structure arguably remain excessively complex. In the near future the pursuit of “high-definition” personalized medicine in psychiatry, is likely to provide an even greater wealth of information regarding factors that influence the dynamic evolution and interaction between symptoms [27]. For instance, experience-sampling techniques and digital phenotyping will allow the monitoring of fluctuations of multiple clinical, environmental or physiological variables in a daily life setting [28]. Network analysis techniques are ideally suited, and are indeed being implemented, to analyze such rich information, which carries tremendous clinical potential [28]. Crucially however, for this complex high-dimensional information to translate into clinical practice, results will not only need to be statistically significant, but should also be intuitively accessible and interpretable. Indeed intuitiveness and communicability remain the main advantages of current diagnostic systems [8]. An ideal framework would need to balance a quantitative low-resolution characterization of the structure of psychopathology similar to factorial analysis, with high-resolution information regarding relevant pathways of interaction between individual symptoms. From this perspective, an approach that seems particularly promising is the use of techniques of dimensionality reduction on graphs, to achieve a topological embedding of individual symptoms that reflects the most salient aspects of the overall network architecture [29]. Employing such topological embedding to a multilayer temporal network of symptoms could offer an intuitive characterization of clinical pathways contributing to the evolution of psychopathology.

The third, and arguably major obstacle to clinical translation, is that psychopathology networks characterize symptom connectivity at the population level, whereas in clinical practice decisions are made on the basis of symptom severity, at the level of the individual. Despite considerable promise, no study to date has, to the best of our knowledge, demonstrated the utility of network approaches in predicting the dynamic development of psychopathology and assist in establishing prognosis. Graph signal processing (GSP) is a relatively novel field of network science that is interested in moving beyond the quantitative characterization of network architecture to model how network architecture effects processes that occur on the network [30]. Similarly, to other branches of network science GSP is devoted to analyzing graphs composed of nodes connected by edges, such as for instance graphs composed of individuals connected by social ties. The unique aspect of GSP is that each node in the graph can be assigned a signal such as for instance the amount of information in a social network, or symptom severity in a psychopathology network. Techniques of GSP can then allow us to study and predict how diffusion of signal on the graph (i.e. diffusion of information among individuals) is influenced by the architecture of connections between nodes (i.e architecture of social bonds) [30]. With regard to psychopathology, techniques of GSP seem extremely attractive to model how dynamic interactions between multiple symptoms will influence heterogeneous clinical evolutions. Specifically, once interactions between symptoms are modeled as a multilayer temporal network, GSP could allow the prediction of how network architecture will influence diffusion of psychopathology across temporal layers at the level of individual patients.

In the present study, we implemented tools of multi-layer network analysis and graph signal processing to characterize and predict clinical pathways of vulnerability to psychopathology in two longitudinal samples of individuals characterized as being at high-risk of developing a psychotic disorder. The first sample is composed of individuals with 22q11.2 Deletion Syndrome, a homogenous genetic disorder, associated with an approximately 30% risk of developing a psychotic disorder [31, 32]. The second sample was composed of individuals at clinical high risk for developing psychosis, recruited from 10 centers internationally in the context of a clinical trial to test efficacy of Poly-Unsaturated-Fatty-Acids (PUFAs) [33]. This first objective was to attempt to provide a quantitative and at the same time intuitive representation of clinical pathways of interaction between symptoms contributing to clinical evolution. The second objective was to use network interactions between symptoms to predict clinical evolution at the level of individual participants. For each section we begin by presenting results in the 22q11DS cohort, followed by results of the replication analysis performed in clinical high-risk individuals.

## Methods

### Sample and Clinical Instruments

#### Primary cohort of individuals with 22q11DS

Individuals with 22q11.2 Deletion Syndrome (22q11DS) were part of a prospective longitudinal study that has been described in several previous publications [34, 35]. Recruitment was performed through patient associations and word of mouth in French and English-speaking European countries. For the present study we included 57 individuals (M/F=26/31), for whom a first psychiatric assessment was available during adolescence (age range at baseline 11.6-18.4, mean 14.4±1.8) along with a second longitudinal assessment on average 3.8±1 years later (age range at follow-up 14.2-24.27, mean 18.25±2.0). Presence of a psychotic disorder at baseline according to DSM-IV-TR criteria was an exclusion criterion.

Psychiatric diagnoses were assessed with the diagnostic Interview for Children and Adolescents-Revised and the psychosis supplement from the Kiddie Schedule for Affective Disorders and Schizophrenia Present and Lifetime Version for individuals below 18 years of age [36, 37]. For adult participants we used the Structured Clinical Interview for DSM-IV Axis I Disorders [38].

To asses sub-threshold positive, negative, disorganized and generalized psychotic symptoms, individuals completed the Structured Interview for Prodromal Syndromes (SIPS) [39]. For a broad characterization of psychopathology, we employed the Brief Psychiatric Rating Scale (BPRS) [40]. To quantify global measures of severity of psychopathology, we employed a combination of the parent-reported versions of the Child Behavior Checklist (CBCL) and Adult Behavior Checklist (ABCL) [41, 42]. Full clinical characterization was performed at both baseline and longitudinal follow-up.

For the primary the construction of multilayer symptom networks we initially consired items of the SIPS and BPRS instruments measured at baseline and longitudinal follow-up. We removed symptoms than had a non-zero score in less that 1% of the sample leading to the exclusion of SIPS grandiosity and BPRS grandiosity scales. This yielded a total of 41 clinical measures available at both baseline and follow up.

#### Replication in individuals at Clinical Ultra High Risk for Psychosis in NEURAPRO cohort

A second cohort of individuals, without a confirmed 22q11.2 Deletion, but meeting criteria for Clinical Ultra High Risk for Psychosis, was recruited in the context of the NEURAPRO clinical trial, designed to test effects of ω-3 Poly-Unsaturated-Fatty-Acid therapy [14, 33]. Individuals were recruited among help-seeking populations in Australia, Singapore, Italy, Germany, Hong Kong, Denmark and Switzerland. Inclusion criteria have been described in detail in previous publications and yielded a total of 304 subjects with a clinical UHR status at baseline. Once included in the study, individuals were randomized to a double-blind 6 month treatment with either ω-3 PUFA or placebo, and were then followed up for further 6 months, yielding a total follow-up period of 12 months [14, 33].

From the original sample of 304 individuals, we excluded 18 subjects with missing assessment of at least 1 item of the Comprehensive Assessment of At-Risk Mental Sate (CAARMS) at baseline, one of which subsequently converted to psychosis. Another 89 subjects were excluded due to missing full characterization at the 12-month follow-up (79 missing CAARMS items, 6 missing BPRS), 19 of which converted to psychosis. This yielded a total of 201 individuals (M/F=98/103) with full clinical characterization at both baseline and 12-month follow-up (age range at baseline: 13.3-37.8 mean 20±4.5)

Psychiatric diagnoses were determined with the Structured Clinical Interview for DSM-IV-TR Axis I Disorders [38]. Sub-threshold positive, negative and generalized psychotic symptoms were evaluated with the Comprehensive Assessment of the At-Risk Mental State [43]. The BPRS was employed for a broad characterization of psychopathology [40] and the Montgomery-Asberg Depression Rating Scale (MADRS) was employed to measure depressive symptoms [44].

Directly comparing network structure across 22q11DS and NEURAPRO cohorts was complicated by the use different SIPS and CAARMS semi-structured clinical interviews across the two samples. Both interviews are designed to assess clinical high risk for developing psychosis, with similar operationalized diagnostic criteria and comparable predictive value [45]. Still, there is no one-to-one correspondence between each item of the two scales. We hence referred to the two manuals two define items that had sufficiently high correspondence across the two instruments. Based on this assessment, we excluded 3 symptoms that were considered to be specific of the SIPS in the 22q11DS and 13 symptoms that were considered as specific on the CAARMS in the NEURAPRO sample. This yielded a total of 37 shared items across the two populations considering both SIPS/CAARMS and BPRS instruments. These items were used to construct longitudinal symptom networks (see Table 1).

**Table 1:**
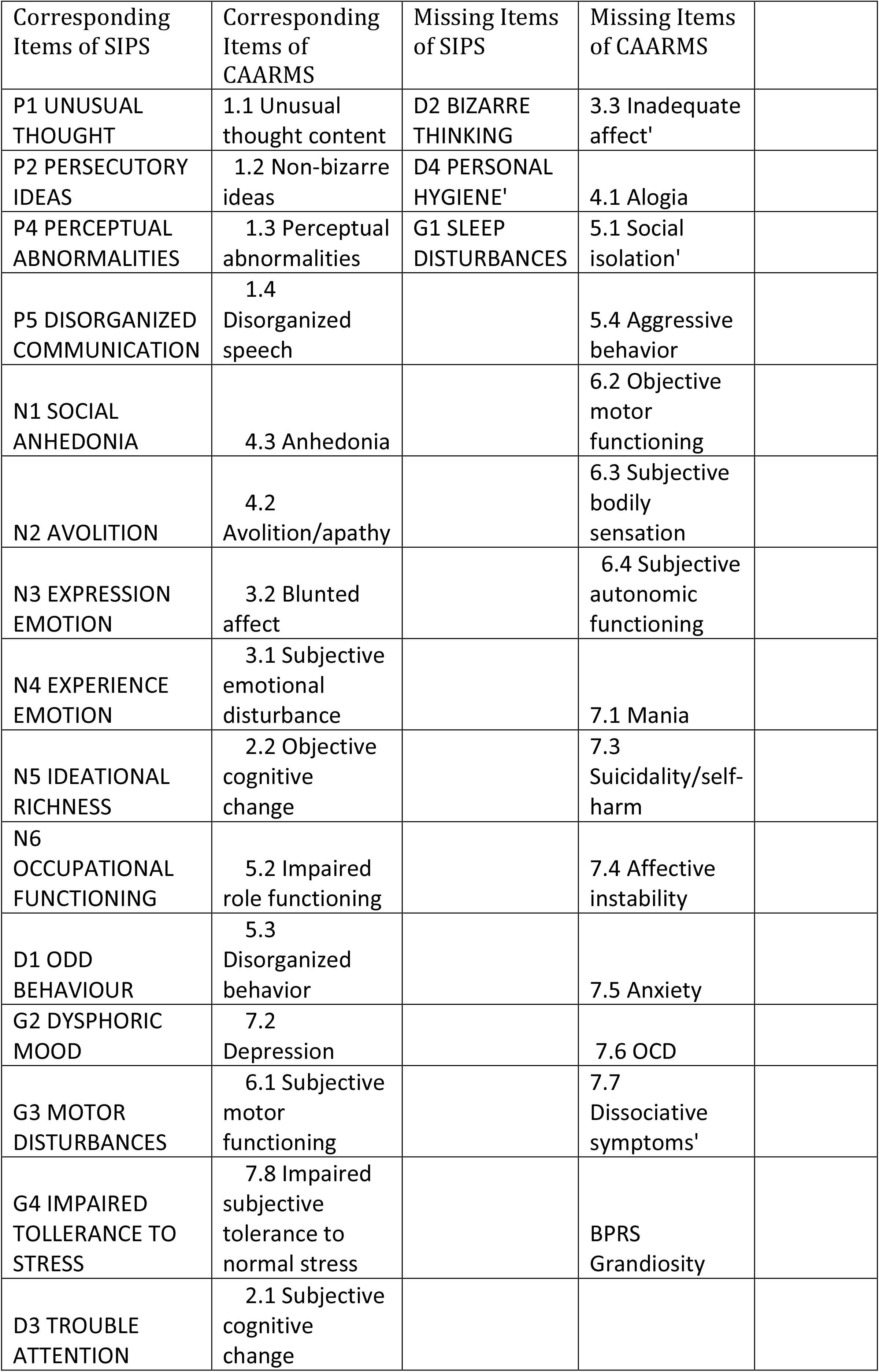
Correspondence of items of SIPS and CAARMS clinical interviews.

## Statistical Analysis Pipeline

### Multilayer symptom networks to define clinical pathways of vulnerability

#### Construction of Multilayer Symptom Networks

Prior to constructing networks, we accounted for the effects of age and sex with linear regression. We then constructed a single multilayer symptom network for each sample, in which each node represented a symptom, and the connecting edge between symptoms was weighted by the Pearson correlation between the two corresponding symptoms across subjects. Graph edges (i.e., correlations) were initially computed cross-sectionally at both baseline and follow up, composing two separate temporal layers. Such separate temporal layers were connected by longitudinal edges estimated from the correlations between symptoms at baseline and symptoms at follow-up, producing a single multi-layer temporal network. Such multilayer network can be expressed in a single adjacency matrix composed of both cross-sectional and longitudinal correlations (See Figure 1). We thresholded the network by considering only correlations survived correction for multiple comparisons with False Discovery at p<0.05.

#### Network topological embedding

In order to investigate main dimensions of variance in the network, we conducted eigendecomposition on the thresholded adjacency matrix representing the multilayer symptom network. Then, network nodes were spatially embedded according to their loading along the two first principal components, yielding a 2-dimensional spatial representation that groups together symptoms that are closely connected in the multilayer network. Eigenvectors of the network provide a *low-dimensional* representation of the main correlation structure between symptoms. However, by simply using these low-dimensional components for the spatial embedding and keeping every symptom as a single node, we retain the *high-dimensional* characterization of the relationships between specific clinical symptoms, both within each time-point and across longitudinal time-points.

#### Graph theory analysis of longitudinal clinical pathways

We investigated several graph theoretical measures to obtain a more quantitative characterization of clinical pathways involved in longitudinal symptom evolution. Specifically, we computed shortest paths connecting each symptom at baseline with each symptom at longitudinal follow-up. We derived a longitudinal betweeness centrality measure by counting the number of longitudinal clinical paths running through each individual symptom. In order to identify symptoms with higher longitudinal betweeness centrality than expected by chance, we constructed 10’000 random networks matched for connectivity by reshuffling edge position. We computed shortest paths connecting symptoms across time in each random network deriving a null-distribution of longitudinal betweeness centrality. P-values for each symptom were computed by estimating the probability of observing a higher betweeness centrality measure than this empirical null-distribution. Further, we used the False Discovery Rate at P<0.05 to correct for multiple comparisons. First, this approach identified symptoms at baseline that over-proportionately mediated effects on symptoms at follow-up. Such longitudinal network hubs at baseline can be conceptualized as *gateways* of psychopathology. Second, our approach identified symptoms at follow-up that were over-proportionately affected and mediated the effects of symptoms at baseline. Such longitudinal network hubs at follow-up can be conceptualized as *funnels* of psychopathology.

### Graph diffusion approach to predict patterns of clinical evolution

Current network approaches to psychopathology have focused on studying the architecture of interactions between psychiatric symptoms mostly by employing techniques of graph-theory [9, 10]. It should however be noted that, in a graph theory framework, symptoms are characterized purely in terms of their connectivity profile with other nodes/symptoms. For network approaches to inform clinical practice at the level of individual patients, symptoms would need to be characterized not only in terms of how they interact with each other, but also in terms of their severity. Indeed, an ideal framework would exploit knowledge of network interactions between symptoms to help predict the evolution of symptom severity across time.

Graph signal processing (GSP) is different from graph theory in that, aside from studying the architecture of network connections, nodes can be assigned a value or signal [30]. Once nodes are assigned a signal in a GSP framework, graph diffusion algorithms have been developed to model how graph architecture influences the propagation of such signal across nodes [30]. An intuitive implementation of this approach is to predict how variations in temperature diffuse over time, across multiple discrete spatial locations. The dynamics of temperature propagation will be determined by the reciprocal distance between spatial locations, with positions that are closer in space having a higher likelihood to influence their neighbor’s temperature, over short periods of time. Graph diffusion addresses this computational problem in a network construct, by modeling discrete spatial locations as nodes in distances as the inverse of connectivity strength between multiple nodes of a network. This then allows predicting how topological network structure influences the dynamics of temperature diffusion.

Graph diffusion approaches are increasingly demonstrating their potential in medical applications. For instance, applying graph diffusion to a multilayer network has been shown to predict the relationship between genetic mutations and tumor samples [46]. Moreover, studies are hinting at the potentials of this approach to model disease progression. Indeed, Raj et al showed that modeling the spread of dementia-related neuro-pathological alterations as a function of the network architecture of long-range axonal fiber bundles reliably predicts the empirically observed patterns of brain atrophy [47]. To the best of our knowledge, graph diffusion approaches have however not yet been employed to the study of psychopathology. Here we propose that applying graph diffusion approaches to multilayer temporal symptoms network can predict the evolution of symptom severity across time, at the level of individual patients.

Our methodological approach, described schematically in Figure 1, began by constructing a multilayer temporal symptom network, excluding the subject for whom we attempted to predict clinical evolution, in a leave-on-out cross validation loop. As previously described the multilayer temporal symptom network was composed both of cross-sectional correlations between symptoms with each time-point and longitudinal correlations between baseline and follow-up symptoms. At the beginning of the diffusion process, symptoms at baseline were assigned a signal that corresponded to the severity that was empirically observed in the excluded subject. Severity of symptoms at follow-up was considered to be unknown and their signal was initially set to 0. The signal corresponding to the empirically observed clinical pattern at baseline was then diffused on the multilayer temporal symptom network, in order to predict symptom severity at follow-up. This is conceptually similar to predicting temperature propagation according to the network structure of distance between nodes.

To predict the spread of symptom severity from baseline to follow-up symptom we employed an iterative finite-difference graph diffusion approach. Compared to simple regression analysis, this approach considers both longitudinal correlations across time-points and cross-sectional correlations between symptoms at follow-up, leading to a progressive evolution and refinement in the predicted symptom pattern. In the example of heat diffusion, the temperature distribution at Time 1 is considered fixed (and therefore re-imposed at each iteration of the algorithm), while the distribution at Time 2 evolves by the diffusion process. For both temperature and psychopathology, the diffusion algorithm will evolve the predicted signal until the system converges towards an equilibrium that minimizes signal change across time, at which point the iterative diffusion will be stopped. The graph diffusion converges to a steady-state solution upon reaching a minimal signal change between iterations that is less than 1e-9. Once such threshold was achieved, the clinical prediction for symptom severity at follow-up was considered to be stable, and the diffusion process was stopped. This process was repeated to predict symptom severity at follow-up for each subject included in the cohort, in a leave-one-out cross-validation loop.

## Results

### Structure of Multilayer Symptom Networks and longitudinal clinical pathways of vulnerability in 22q11DS

Topological embedding of symptoms yielded a strong negative correlation between the Euclidian distance separating symptoms and the empirically observed correlation strength (R=-0.465, P<0.0001, observed not only for cross-sectional associations between symptoms at baseline (R=-0.354, P<0.0001) or at follow-up (R=-0.473, P<0.0001), but also for longitudinal associations between symptoms at baseline and symptoms at follow-up (R=-0.365, P<0.0001) See Figure 2C). These results suggest that an easily interpretable low-dimensional embedding can offer a good approximation of the structure of the multilayer symptoms network.

**Figure 2.**
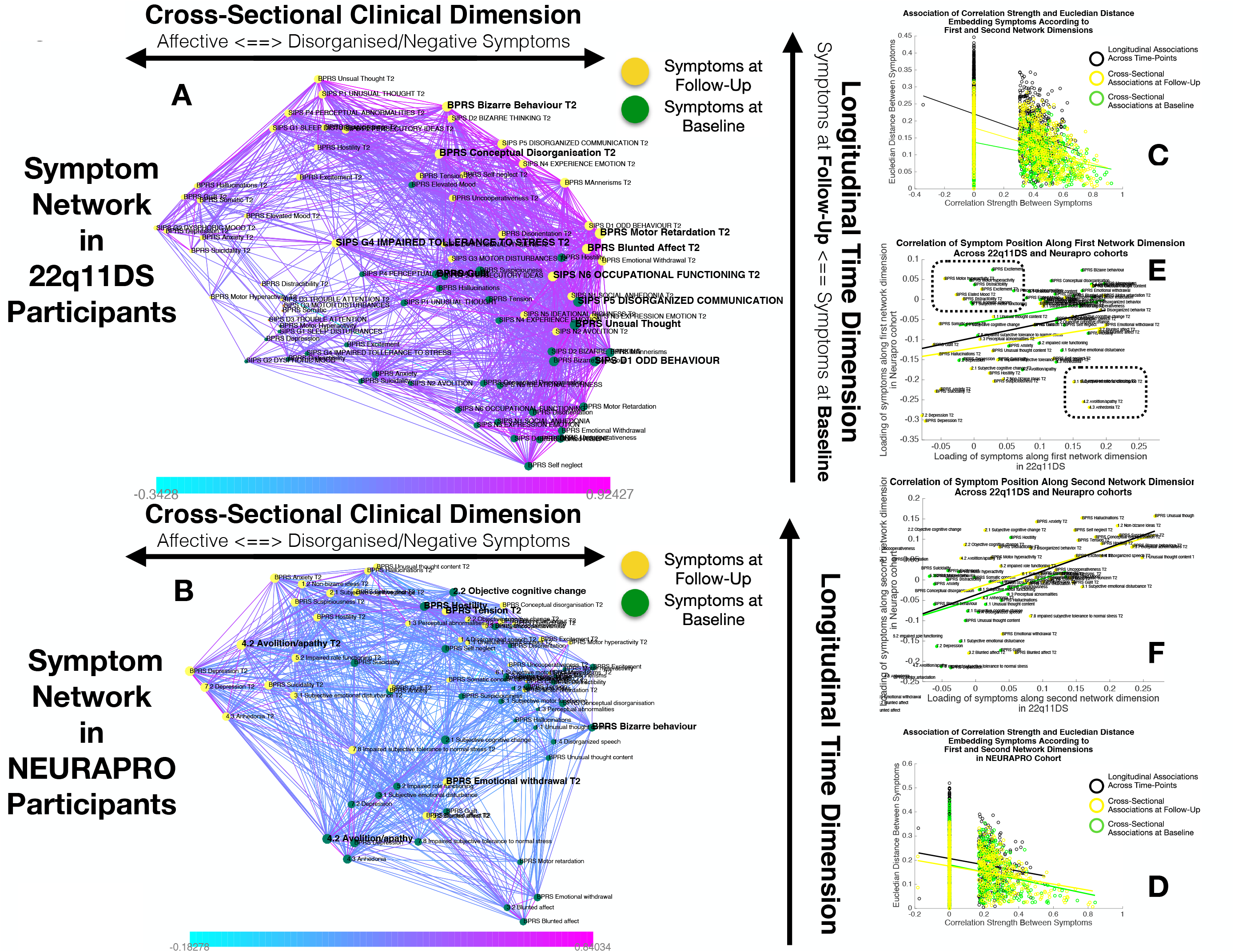
**A-B:** Structure of longitudinal symptom networks in 22q11DS sample**: A** and NEURAPRO sample: **B**. Spatial embedding of symptoms according to network dimensions derived from singular value decomposition. The first dimension is plotted along the horizontal X axis whereas the second dimension is plotted along the vertical Y axis. Lines connecting symptoms represent correlations that survive correction for multiple comparisons at P<0.05 color-coded according to correlation strength. Symptoms at baseline are displayed in green and symptoms at follow-up are displayed in yellow. Size of nodes is scaled according to mean connectivity strength of each symptoms. Symptoms that present a higher than random centrality in mediating clinical pathways going from baseline to follow-up are displayed in bold. **C-D:** Association of Euclidian distance between symptom after spatial embedding according to first and second network dimensions and empirically observed correlation strength between symptoms in 22q11DS sample: **C** and NEURAPRO sample: **D**. Cross-sectional associations between symptoms at baseline are displayed in green and between symptoms at follow-up are displayed in yellow. Longitudinal association between symptoms at baseline and symptoms at follow-up are displayed in black. **E:** Association between position of symptoms according to the first network dimension across 22q11DS and NEURAPRO cohorts. Two clusters of symptoms that contribute negatively to the correlation between structures of symptom networks across the two cohorts, suggesting a different pattern of correlation with other forms of psychopathology, are circled. **F:** Association between position of symptoms according to the second network dimension across 22q11DS and NEURAPRO cohorts.

The first network dimension, plotted along the horizontal axis in Figure 2A, mainly captured the structure of cross-sectional correlations between symptoms within each time-point. Symptoms located on the right side of the graph mainly captured disorganization and thought disorder including SIPS Odd Behavior and Disorganized communication and BPRS Bizarre Behavior, Mannerism and Unusual Thought Content. Negative symptoms were mostly located on the right-side of the graph, near disorganization symptoms. The opposite left side of the graph, was on the other hand, populated by symptoms of affective dysregulation, including SIPS Dysphonic Mood and Reduced Tolerance to Normal Stress and BPRS Depression and Anxiety. Symptoms of attention deficit hyperactivity disorder (ADHD) including SIPS Trouble with Attention, BPRS Distractibility and Motor Hyperactivity, were located on the left side of the graph near affective disturbances. Positive symptoms had an intermediate position along the first dimensions, with SIPS Perceptual Abnormalities and BPRS Hallucinations being closer to affective and ADHD symptoms, whereas SIPS and BPRS Thought Disorder were closer to negative and disorganized symptoms. Loading of symptoms along this first eigen-vector was highly correlated across time-points (R=0.7, p<0.0001), pointing an overall stability in cross-sectional structure of the symptom network over time, along an affective to negative/disorganized dimension.

The second dimension was plotted along the vertical Y axis and predominantly captured the temporal aspect, with symptoms at baseline located at the bottom of the graph and symptoms at follow-up being located at the top of the graph. Importantly, aside from an overall distinction of symptoms across-time points, we observed a significant variation along the time dimension between symptoms measured within each time-point, which captured the differential propensity of symptoms to influence one another over time. Indeed, we observed an opposite association across the two time-points between loading of symptoms according to second time dimension and the mean strength of longitudinal correlations between symptoms at baseline and symptoms at follow-up (at baseline R=0.3,P=0.05, at follow-up R=-0.22,P=0.16, P of difference = 0.0094; See Supplementary Figure 1). In this perspective, symptoms that that were higher than average at baseline can be considered more highly predictive of psychopathology at follow-up. On the opposite, symptoms that were located lower that the rest at follow-up, were more directly influenced by prior psychopathology at baseline. This representation offered an intuitive characterization of the relationship between symptoms over time.

Subsequently we were interested in highlighting clinical pathways involving individual symptoms that played a particularly prominent role in disease progression. Our approach based on graph theory identified 4 symptoms at baseline that disproportionately affected clinical symptom pattern at follow-up, and that can be conceptualized as gateways of psychopathology (shown in bold in Figure 2A. Network embedding presented before provided an intuitive characterization of the different longitudinal clinical pathways affecting such gateway symptoms. The first three symptoms were located on the right side of the graph and mainly captured thought disorder and disorganization including SIPS Disorganized Communication, SIPS Odd Behaviour and BPRS unusual thought. Disorganization symptoms, such as SIPS Odd Behavior, acted as a gateway by broadly mediating the effects negative symptoms at baseline on both disorganized and negative symptoms at follow-up (See Figure 3D). A fourth gateway symptoms was represented by BPRS guilt. BPRS guilt was located closer the left side of the graphs and acted as a gateway by broadly mediated the of effects affective symptoms at baseline on both affective and thought disturbance symptoms at follow-up (See Figure 3C).

**Figure 3.**
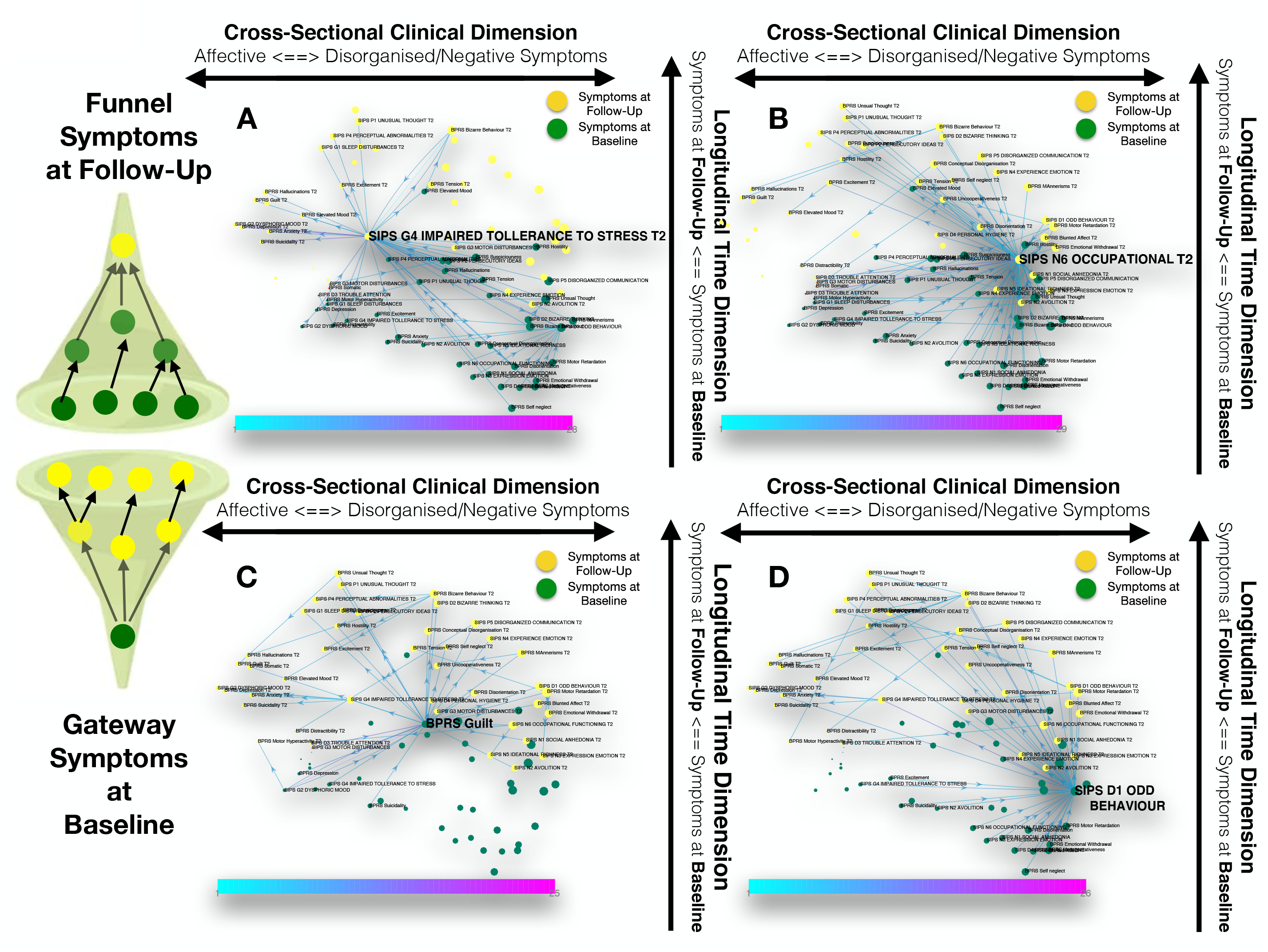
Longitudinal clinical pathways running through gateway symptoms at baseline (**C-D**) and funnel symptoms at follow-up (**A-B)** in 22q11DS sample. **A:** Impaired tolerance to daily stress at follow-up acts as a funnel by broadly mediating the effects of baseline of thought disturbances on follow-up affective symptoms and of baseline affective symptoms on follow-up thought disturbances. **B:** Reduced occupational functioning at follow-up, acted as a funnel by broadly mediating the effects of negative, disorganized symptoms and ADHD symptoms at baseline on the persistence of negative and disorganized symptoms at follow-up. **C:** BPRS guilt at baseline acts as a gateway by mediating the of effects affective symptoms at baseline on both affective and thought disturbance symptoms at follow-up. **D:** SIPS Odd Behavior acted as a gateway by broadly mediating the effects negative symptoms at baseline on both disorganized and negative symptoms at follow-up.

Our approach also identified 6 symptoms at follow-up, that were broadly affected by psychopathology at baseline, and that can hence be conceptualized as funnels of psychopathology. Two of these funnel symptoms were captured disorganization and were represented by bizarre behavior and conceptual disorganization, and mostly mediated the effects of priori disorganization symptoms. Two more were represented by negative symptoms such as BPRS blunted affect and SIPS occupational functioning, which were located on the right side of the graph and appeared to importantly mediated the effects of negative and disorganized symptoms and ADHD symptoms at baseline on the persistence of negative and disorganized symptoms at follow-up (See Figure 3B). A final funnel was represented by SIPS reduced tolerance to normal stress, which located left side of the graph appeared important in mediating the effects of baseline of thought disturbances on follow-up affective symptoms and of baseline affective symptoms on follow-up thought disturbances (See Figure 3A).

### Structure of Multilayer Symptom Networks and longitudinal clinical pathways of vulnerability in NEURAPRO sample

While the variance explained by the first two dimensions was lower in the NEURAPRO sample, we still observed a significant negative correlation between the Euclidian distance separating symptoms and the empirically observed correlation strength (R=-0.249, P<0.000; See Figure 2D), observed for both cross-sectional associations between symptoms at baseline (R=-0.249, P<0.0001) or between symptoms at follow-up (R=-0.238, P<0.0001), and for longitudinal associations between symptoms at baseline and symptoms at follow-up (R=-0.135, P<0.0001). This suggests that spatial embedding of symptoms according to just the two main eigenvectors still offered a meaningful characterization of the interaction between individual symptoms

Similarly to results in 22q11DS, the first dimension mainly captured variance between cross-sectional correlations within each time-point (See Figure 2B). Symptoms located on the right side of the graph were mostly composed of positive and disorganized symptoms, including Bizarre Behavior, Unusual Thought and Hallucinations measured with both BRPS and CAARMS. The left side of the graph was mostly populated by symptoms of affective disturbances, including Depression and Anxiety, BPRS Guilt and CAARMS Subjective Reduced Tolerance to Daily Stressors. Negative symptoms could be divided in two sub-groups according to their loading along the first dimension. Specifically, symptoms of reduced emotional expressiveness, such as Blunted Affect, BPRS Emotional withdraw, and CAARMS Anhedonia were located on the right side of the graph, closer to disorganized symptoms. On the opposite symptoms of reduced motivational drive such CAARMS Avolition/Apathy and Impaired Role Functioning were located closer to the left side of the graph and closer to anxiety/depressive symptoms. Interestingly we observed a significant positive correlation between the loading of symptoms along the first “cross-sectional” dimension (R=0.299, P=0.008) across 22q11DS and NEURAPRO samples (See Figure 2E). This would suggest a similar structure of cross-sectional psychopathology across 22q11DS and NEURAPRO samples, mainly reflecting an overall distinction of affective and negative-disorganized symptoms. Still, in the context of an overall similar network structure two groups of symptoms appeared to cluster differently in the networks of the two samples (Circles in Figure 2E). In particular, symptoms of ADHD, including BPRS motor hyperactivity and distractibility, were in proximity to affective symptoms in 22q11DS, whereas they were closer to symptoms of thought disorder in the NEURAPRO sample. Moreover, a sub-group of negative symptoms including experience of emotion, avolition, social anhedonia and occupational functioning were located closer to other negative and disorganized symptoms in the 22q11DS cohort whereas they clustered closer to depressive and affective symptoms in the NEURAPRO sample.

The second network dimension mainly captured the dimension of time, with symptoms at baseline mainly located at the bottom of the graph and symptoms at follow-up mainly located at the top of the graph. Similarly, what was found in the 22q11DS cohort there was significant variance within symptoms at each time-point along this time dimension. Interestingly the correlation of symptom loading across samples was even stronger along this second longitudinal dimension (R=0.56, P<0.0001 See Figure 2F). suggesting that the relative predictive value of symptoms at baseline in influencing symptoms at follow-up, and the relative tendency of symptoms at follow-up to be influenced by prior psychopathology, is similar across the two clinical populations.

Our approach identified 3 baseline symptoms that presented disproportionately high centrality in mediating clinical patterns at follow-up displayed in Figure 4. In particular BPRS bizarre behavior was located on right-side of the graph and appeared to broadly affect negative and disorganized symptoms at follow-up. Moreover, bizarre behavior indirectly affected subsequent affective disturbances p through the mediating role of emotional-withdrawal at follow-up. BPRS hostility was also located in proximity to negative and disorganized symptoms at baseline and appeared central in mediating their effects on subsequent symptoms of mood disturbance. Finally, avolition-apathy was located on the left side of the graph and was directly associated with subsequent affective symptoms and indirectly associated with negative and disorganized symptoms, through the mediating role of persistent avolition-apathy at follow-up. Indeed avolition-apathy at follow-up was also highlighted as a key funnel symptoms that broadly mediated the effects of baseline avolition and affective disturbances on subsequent psychopathology. Despite an overall similar network structure appeared similar in 22q11DS and NEURAPRO cohorts, we did not observe a significant association in measures of longitudinal betweeness centrality (R=-0.03, P=0.77). These results suggest that specificities exist in the role of individual symptoms in contributing to the evolution of psychopathology, across the two samples.

**Figure 4.**
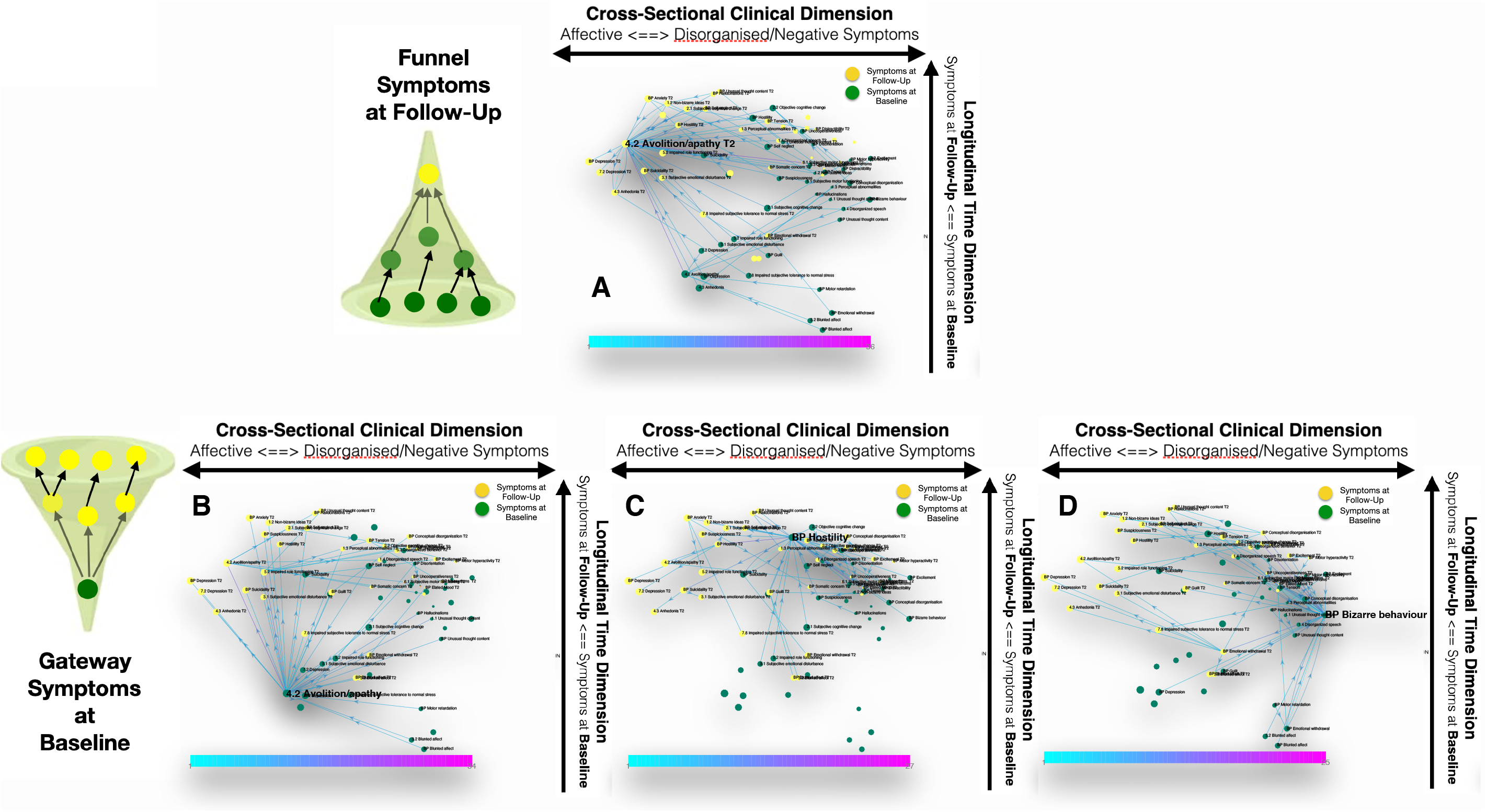
Longitudinal clinical pathways running through gateway symptoms at baseline (**B-D**) and funnel symptoms at follow-up (**A)** in NEURAPRO sample. **A:** avolition-apathy at follow-up was also highlighted as a key funnel symptoms that broadly mediated the effects of baseline avolition and affective disturbances on subsequent psychopathology.**B:** Avolition-apathy was located on the left side of the graph and was directly associated with subsequent affective symptoms and indirectly associated with negative and disorganized symptoms, through the mediating role of persistent avolition-apathy at follow-up. **C:** BPRS hostility was also located in proximity to negative and disorganized symptoms at baseline and appeared central in mediating their effects on subsequent symptoms of mood disturbance. **D:** BPRS bizarre behavior was located on right-side of the graph and appeared to broadly affect negative and disorganized symptoms at follow-up. Moreover, bizarre behavior indirectly affected subsequent affective disturbances p through the mediating role of emotional-withdrawal at follow-up.

### Graph diffusion approach to predict patterns of clinical evolution

#### Evaluation of prediction accuracy in 22q11DS and NEURAPRO cohorts

Our primary objective was to predict the multivariate patterns of symptoms included in the SIPS and CAARMS clinical interviews, designed to assess vulnerability to psychosis. We started by predicting severity of SIPS and CAARMS items at follow-up using items of SIPS and CAARMS at baseline. Subsequently, we estimated the added benefit of considering additional clinical instruments at baseline.

Simply correlating severity at baseline against severity at follow-up for each symptom across subjects, revealed a positive significant correlation both in the 22q11DS cohort for 14/18 symptoms being tested (R=0.37±0.12) and in the Neurapro cohor for 27/28 symptoms (R=0.31±0.1; See Figure 5A and 6A, respectively). While not surprising, these results suggest that simply considering a patient as clinically stable across time-points provides a highly non-random estimate of the clinical pattern at follow-up. We computed the mean squared change in symptom severity across time-points across all subjects and symptoms. This Mean-Squared-Error is a measure of prediction accuracy achieved by simply considering clinical stability across time-points, that we used as a baseline against which we tested the performance of our graph-diffusion based prediction approach.

**Figure 5.**
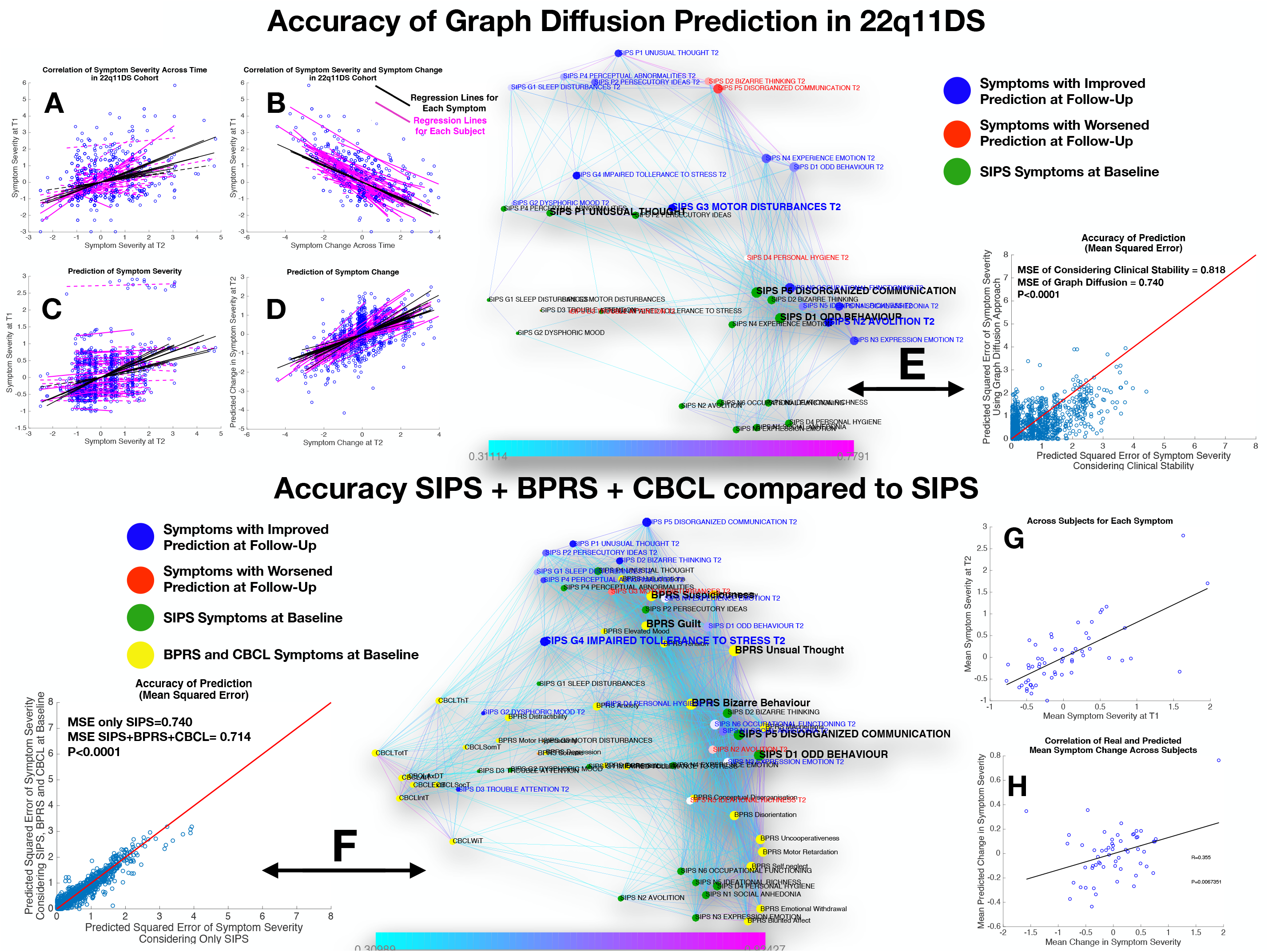
Performance of graph diffusion approach in predicting patters of SIPS psychopathology at longitudinal follow-up in 22q11DS sample. **A:** Correlation of symptom severity across time-points for all symptoms across participants. Regression lines for symptoms are displayed in black with dashed lines indicating correlations that are not significant at P<0.05. Regression lines for subject are displayed in purple with dashed lines indicating correlations that are not significant at P<0.05. **B:** Correlation between symptom severity at baseline and change in symptom severity between baseline and follow up for all symptoms across all participants. **C:** Correlation between real and predicted symptom severity at follow-up. **D:** Correlation between real and predicted symptom change between baseline and follow-up. **E:** Comparison of accuracy in predicting SIPS at follow-up, between considering clinical stability and graph diffusion approach using SIPS at baseline. Symptoms are spatially embedded according to two main network dimensions derived from SVD. Symptoms at baseline are displayed in green. Symptom at follow-up are color coded according to prediction accuracy of graph diffusion compared to considering clinical stability, with blue symptoms having higher accuracy using graph diffusion and red symptoms having worsened accuracy. **F:** Prediction accuracy of considering the combination of SIPS, BPRS and CBCL at baseline compared to using only items of the SIPS. Symptoms are spatially embedded according to two main network dimensions derived from SVD. Symptoms of the SIPS at baseline are displayed in green. Items of additional clinical instruments are displayed in yellow. Symptom at follow-up are color coded according to prediction accuracy of considering an additional clinical instrument compared to accuracy achieved by using only items of the SIPS, with blue symptoms having higher accuracy and red symptoms having worsened accuracy. **G:** Correlation between mean symptom severity at follow-up and mean predicted symptom severity at follow-up. **H:** Correlation between mean symptom change across time-points and mean predicted symptom change.

A perhaps less intuitive observation was that change in symptom severity between baseline and follow-up was strongly negatively correlated with symptom severity at baseline for all symptoms being tested in both the 22q11DS (R=-0.65±0.17) and Neurapro cohorts (R=-0.63±0.25), suggesting the existence of a phenomenon of regression to the mean (See Figure 5B and 6B, respectively).

#### Performance of prediction in 22q11DS sample

Considering only the SIPS subscale at baseline yielded a significant prediction of SIPS symptom severity at follow-up, as revealed by a strongly significant correlation between actual and predicted symptom severity (R=0.40, P<0.00001) across all items and subjects, that remained significant when averaging mean and predicted symptom severity in each subject (R=0.64, P<0.00001; see Figure 5C and 4E). Interestingly, the correlation between empirical and predictive values was even stronger when considering symptom change across the two time points for all symptoms and subjects (R=0.57, P<0.00001). However, when averaging symptom change in each subject, we did not observe a significant correlation between observed and predicted values (R=-0,22, P=0.08). In other terms, the algorithm predicted both mean and specific symptoms severity at follow-up and specific change in symptom severity, while it failed to predict the mean change in symptom severity (see Figure 5F).

Importantly, prediction accuracy of graph diffusion was significantly higher that simply considering clinical stability (MSE of clinical stability = 0.818 ± 0.82, MSE of graph diffusion = 0.74± 0.62, p<0.00001; see Figure 3G). Exploring the distribution in the difference of prediction accuracy across symptoms revealed that accuracy of graph diffusion was higher for all symptoms except personal hygiene, bizarre thinking, disorganized communication and trouble with attention (see Figure 5H).

Next, we were interested in assessing the added value of considering additional clinical instruments at baseline. Adding the BPRS evaluation at baseline provided a small but significant improvement in SIPS prediction at follow-up (MSE of SIPS = = 0.74± 0.62, MSE of SIPS + BPRS = 0.72± 0.60 P=0.03), whereas adding the CBCL at baseline did not significantly improve the accuracy of symptom prediction at follow-up (MSE of SIPS= 0.74± 0.62, MSE of SIPS + CBCL= 0.73± 0.60 P=0.316). However, when considering the combination of adding CBCL + BPRS, this yielded a strong increase in prediction accuracy that was highly significant compared to considering only the SIPS (MSE of SIPS= 0.74± 0.62, MSE of SIPS + BPRS+ CBCL = 0.716±0.59 P<0.00001) or separately adding BPRS (P<0.00001), or CBCL (p<0.00001) (see Figure 3C). Moreover, in addition to significantly predict mean symptom severity (R=0.69, P<0.00001), relative symptom severity (R=0.45, P<0.00001), and relative symptom change (R=0.61, P<0.00001), adding BPRS and CBCL significantly predicted mean change in SIPS symptom severity over time (R=0.35, P=0.006; see Figures 5 C and D).

These results point to a synergism of BPRS and CBCL in predicting clinical patterns of the SIPS at follow-up. Interestingly this synergism was visually apparent from the position of the items of the two instruments within the structure of the longitudinal symptom graph. Indeed, while items of the CBCL clustered on the left side of the graph in proximity to affective and ADHD symptoms, most items of the BPRS were located on the right side of the graph in proximity to thought disorder and negative symptoms.

#### Performance of prediction in NEURAPRO sample

Similarly, to what was observed in the 22q11DS, the graph diffusion approach yielded a significant prediction of patterns of symptom severity at follow-up, with an average correlation between real and predicted symptom severity across all subjects (R=0.26, P<0.0001; see Figure 6C). Correlation was stronger between real and predicted symptom change between baseline and follow-up (R=0.54, P<0.0001; see Figure 6D).When averaging severity across symptoms for each subject we observed a significant correlation between mean and predicted symptoms severity (R=0.51,P<0.0001) but not between mean and predicted change in symptom severity (R=0.04,P=0.56), similar to what was observed in the 22q11DS cohort (see Figures 6 E and F).We hence compared prediction accuracy of the graph diffusion approach against that of simply considering clinical stability across time. As in 22q11DS, this analysis revealed that MSE of the graph diffusion approach was significantly lower that simply considering clinical stability (MSE of clinical stability = 0.871 ± 0.79, MSE of graph diffusion = 0.734± 0.67, p<0.00001; see Figure 6G). Indeed, accuracy of prediction was higher for all items of the CAARMS except inadequate affect, objective motor functioning and mannerism (see Figure 6H).

**Figure 6.**
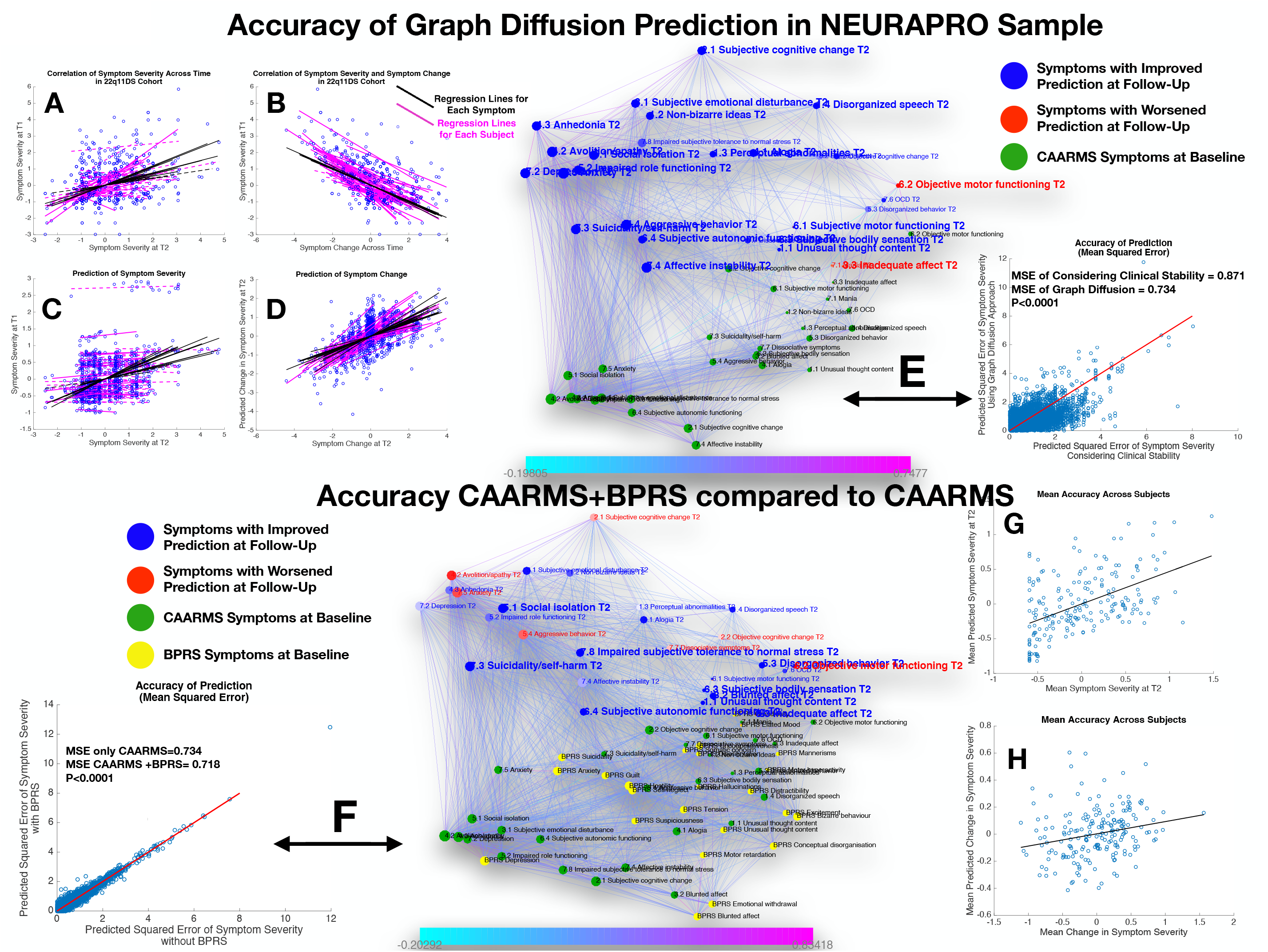
Performance of graph diffusion approach in predicting patters of CAARMS psychopathology at longitudinal follow-up in NEURAPRO sample. **A:** Correlation of symptom severity across time-points for all symptoms across participants. Regression lines for symptoms are displayed in black with dashed lines indicating correlations that are not significant at P<0.05. Regression lines for subject are displayed in purple with dashed lines indicating correlations that are not significant at P<0.05. **B:** Correlation between symptom severity at baseline and change in symptom severity between baseline and follow up for all symptoms across all participants. **C:** Correlation between real and predicted symptom severity at follow-up. **D:** Correlation between real and predicted symptom change between baseline and follow-up. **E:** Comparison of accuracy in predicting CAARMS at follow-up, between considering clinical stability and graph diffusion approach using CAARMS at baseline. Symptoms are spatially embedded according to two main network dimensions derived from SVD. Symptoms at baseline are displayed in green. Symptom at follow-up are color coded according to prediction accuracy of graph diffusion compared to considering clinical stability, with blue symptoms having higher accuracy using graph diffusion and red symptoms having worsened accuracy. **F:** Prediction accuracy of considering the combination of CAARMS and BPRS at baseline compared to using only items of the CAARMS. Symptoms are spatially embedded according to two main network dimensions derived from SVD. Symptoms of the CAARMS at baseline are displayed in green. Items of BPRS at baseline are displayed in yellow. Symptom of CAARMS at follow-up are color coded according to prediction accuracy of considering an additional clinical instrument compared to accuracy achieved by using only items of the CAARMS, with blue symptoms having higher accuracy and red symptoms having worsened accuracy. **G:** Correlation between mean symptom severity at follow-up and mean predicted symptom severity at follow-up. **H:** Correlation between mean symptom change across time-points and mean predicted symptom change.

Next, we estimated the additive predictive value of considering additional clinical instruments at baseline. Adding the BPRS at baseline provided a strong improvement in the prediction of CAARMS items at follow-up (MSE of CAARMS = 0.734± 0.67, MSE of CAARMS+ BPRS = 0.718±0.66 p<0.0001; see Figure 7C). Moreover in addition to significantly predict mean symptom severity (R=0.50,P<0.0001), relative symptom severity (R=0.25, P<0.00001), and relative symptom change (R=0.57, P<0.00001), adding BPRS significantly predicted the mean change in CAARMS symptom severity over time (R=0.29, P=0.001; see Figures 67D and E). On the other hand, adding MADRS scores did not significantly improve average prediction accuracy (MSE of CAARMS = 0.734± 0.67, MSE of CAARMS+ MADRS = 0.734±0.67, P=0.589; see Figure 7A). Moreover, considering the addition of BRPS + MADRS worsened the accuracy of prediction compared to the combination of CAARMS and BPRS (MSE of CAARMS+ BPRS 0.718±0.66, MSE of CAARMS+ MADRS = 0.723±0.67 p<0.0001). See Figure 7B. The lack of added predictive value of the MADRS to the CAARMS would have been predicted from the position of the MADRS items at baseline within the structure of the longitudinal symptom network. Indeed, although MADRS items were located on left “affective” side of the graph, they were located lower along the time dimension that corresponds to affective items of the CAARMS. This would suggest that the CAARMS characterization of affective dysregulation at baseline is sufficient and indeed superior to MADRS items in terms of predicting CAARMS psychopathology at follow-up.

## Discussion

Current clinical approaches to tackle the complexity of mental health disturbances have almost invariably merged together clinical manifestations that often co-occur across participants. However, especially in the earliest stages of psychopathology, merging clinical manifestations may hinder our understanding of pathways of interaction between individual symptoms, which in turn may be relevant for predicting prognosis or planning treatment strategies. Network approaches to psychopathology represent a promising framework to model complex disease pathways between individual symptoms, but two main factors may have to date limited their clinical translation.

The first limitation refers to the insufficient intuitiveness and interpretability of results of current network analyses. We argue that such insufficient interpretability is the combined result of the application of network approaches to cross-sectional data, together with the excessive complexity of resulting symptom networks. In the present manuscript we propose a methodological approach based on multi-layer network analysis that offers an intuitive and quantitative and quantitative characterization of clinical pathways of interaction between symptoms over time.

The second main limitation is that current network approaches characterize symptoms exclusively in terms of their reciprocal interactions, which are estimated at the level of a population. Clinical practice on the other hand entails making predictions about symptom severity at the level of the individual. Here we propose that a network approach inspired by graph signal processing can allow to combine information regarding symptom *connectivity* and *severity* allowing to predict multivariate patterns of clinical evolution at the level of individual participants.

We test our approach in two independent samples of individuals at risk for the developing psychosis.

### Temporal Multilayer Symptom Network approach to characterize clinical pathways of vulnerability to psychopathology

A prerequisite for interpreting the role of specific symptoms is having a broad characterization of the overall structure of psychopathology, similar to seeing the outline of the forest before focusing on the trees. In both samples, the first network component captured to the overall cross-sectional structure of relationships between symptoms, largely reflected a distinction between affective vs negative-disorganized psychopathology. Such cross-sectional structure was conserved both across longitudinal visits and across samples and is consistent with results of classical factorial analysis in both high-risk populations and schizophrenia [48-51] This would suggest that overall network architecture reflects broad clinical patterns observed in clinical practice, and confirms the previously hypothesized distinction between affective and negative/disorganized dimensions of vulnerability to psychosis [52, 53]. It worth noting however, that compared to our approach, factorial analysis separates symptoms that considered to be expression of distinct underlying latent variables. Therefore, by design, factorial analysis sacrifices information residing in the structure of correlations observed within and a cross large-scale dimensions [10]. By comparison, spatial embedding of individual symptoms captures the relationship between large-scale symptoms, such as the relative proximity of negative and disorganized dimensions, as well as the potential existence of relevant sub-clusters within large-scale dimensions. For instance, in both samples avolition was located closer to affective and depressive symptoms compared to symptoms of reduced emotional expressiveness, which is in agreement with evidence of the existence of sub-dimensions within negative symptoms [54].

Aside from to the structure of cross-sectional psychopathology, the key advantage of the MTSN approach is the ability to capture pathways of longitudinal interactions between symptoms. Indeed, despite the inherent dynamic nature of the “Network Theory of Psychopathology” most network analyses are conducted on cross-sectional data, hence lacking the essential dimension of time. In our approach, the time dimension was intuitively captured in the second network component plotted along the vertical axis, with symptoms at baseline located at the bottom of the graph and symptoms at follow-up located at the top. Euclidian distance between symptoms offers therefore an intuitive characterization of the propensity of different clinical manifestations to influence one-another across longitudinal assessments. For instance, according to the first cross-sectional dimension negative symptoms of reduced emotional expressiveness were located in proximity to symptoms of conceptual disorganization and thought disturbances. However, in both samples the second time dimension clearly distinguished between the two forms of psychopathology, with baseline symptoms of thought disturbance located much closer to psychopathology at follow-up compared to reduced emotional expressiveness. This finding would suggest that symptoms of reduced emotional expressiveness develop as a consequence of prior thought disturbance and disorganization, and have hence a less active role in influencing subsequent psychopathology. Such interpretation is consistent with literature on basic symptoms of psychosis that suggests that subclinical subjectively experienced thought disturbances lie at the core of the phenomenology of the disorder and play an active role in influencing clinical evolution and particularly negative symptoms [55].

One of the main challenges in developmental and early-intervention psychiatry is the growing realization that early clinical manifestations of psychopathology are largely not specific to a single clinical outcome. We propose that cross-diagnostic clinical evolutions may related to specific mechanisms that act as developmental cross-roads in evolution of psychopathology. In particular some clinical manifestations may broadly increase risk for subsequent psychopathology, while others may be broadly affect different forms of prior psychopathology. Targeting such symptoms where the “flow” of psychopathology either broadens or narrows could be particularly effective in preventing deleterious clinical outcomes. The MLSN is ideally suited to identify such *gateways* and *funnels* of psychopathology, offering an intuitive characterization of longitudinal clinical pathways over time. For instance, in both samples our analysis confirmed that sub-threshold manifestation of though disturbance, acted as gateways, broadly increasing the risk for subsequent psychopathology. On the opposite, negative symptoms such as blunted affect and occupational functioning in 22q11DS or avolition-apathy in the NEURAPRO sample acted as funnels that were broadly passively influenced by prior psychopathology. Moreover, some symptoms appeared to act as crossroads bridging across the affective to negative-disorganized dimensions over time. For instance, Hostility in the NEURAPRO sample was associated with thought disturbances at baseline but increased the risk for developing affective symptoms at follow-up. On the opposite, guilt in 22q11DS was associated with affective symptoms at baseline but increased the risk for both affective symptoms at thought disturbances at follow-up. Finally, particularly in the 22q11DS sample our results pointed to an important role of reduced tolerance stress at follow-up, in firstly mediating the effects of prior effective disturbances on subsequent psychotic symptoms. These findings are strongly reminiscent of the reduced tolerance to stress in the “affective pathway” to psychosis initially proposed by Myin-Germeys and Van Os [56]. Moreover, our findings also suggest that reduced tolerance to stress may partially mediate the effects of prior thought disorder on the subsequent development of affective disturbances. The prominent role of this pathways in 22q11DS may be related to recent evidence of dysregulation of the dysregulation of the Hypothalamus-Pituitary-Adrenal-Axis [57] and heightened vulnerability to environmental stress in this population [58].

Altogether results both in 22q11DS and NEURAPRO cohorts highlight the potentialities of an approach based on multilayer temporal network analysis to provide and intuitive and quantitative characterization of clinical pathways contributing to heterogenous clinical evolutions in the early stages of psychopathology.

### Predicting clinical evolution of individual patients through multi-layer graph diffusion

Aside from shedding light on underlying disease mechanisms, a major appeal of understanding pathways of interaction between symptoms, is in assisting in establishing prognosis. Still, current network approaches to psychopathology characterize symptoms exclusively in terms of their reciprocal connectivity profile, sacrificing information regarding symptom severity in individual participants. The unique feature of Graph Signal Processing (GSP) is that network nodes are characterized not only in terms of connectivity, but can also be assigned a value or signal. For instance, in our GSP approach baseline symptoms were assigned a signal that corresponded to their observed severity in a particular subject. For each subject we then predicted evolution of psychopathology by modeling the diffusion of symptom severity from baseline to follow-up symptoms, as function of the structure of the multi-layer temporal symptom network (See Figure 2).

To the best of our knowledge, our results are the first to demonstrate the potentialities of a purely network based graph diffusion approach in predicting multivariate patterns of clinical evolution at the level of individual participants. Importantly, in both samples, prediction accuracy was significantly higher than simply considering clinical stability across time in both 22q11DS and NEURAPRO samples. It has been argued that an excessive focus on a single dichotomous clinical outcome such as conversion to psychosis might represent a major limit of the current UHR framework [59]. Indeed the presence of a UHR status increases the likelihood of developing a range of psychopathological outcomes that have the potential to negatively influence an individual’s functional outcome [2, 13, 59, 60]. The potential for diverse psychiatric outcomes is also well described in individuals carrying genetic risk for psychosis [61] including in 22q11DS [32]. Indeed besides a 30% risk of developing a psychotic disorder individuals with 22q11DS present a 30% likelihood of presenting an anxiety disorder, a 30% likelihood of being diagnosed with ADHD and a 20% risk of developing a mood disorder by adulthood, all of which can negatively affect quality of life [32]. A significant advantage of a network-based graph diffusion approach is that clinical prediction is performed at the level of individual symptoms with the potential of describing mixed and heterogeneous clinical evolutions.

Aside from flexibility in terms of considering clinical outcome the network-based graph diffusion approach is also flexible in terms of integrating multiple predictors at baseline. Indeed, results in both samples suggest that a broad clinical characterization at baseline, that goes beyond considering associations between homologous forms of psychopathology, can improve prediction of clinical outcome at follow-up. Specifically, in the NEURAPRO sample prediction accuracy of CAARMS items was improved when considering a combination of CAARMS and BPRS at baseline whereas in the 22q11DS sample prediction accuracy was strongly improved when adding a combination of BPRS and CBCL at baseline. Interestingly, network dimensionality reduction offered an intuitive appreciation of reasons underlying the value of additional clinical instruments in improving prediction accuracy. Indeed, the synergism of CBCL and BPRS was related to the fact that two instruments appeared to capture opposite facets of psychopathology, with CBCL assisting prediction of affective and ADHD symptoms while most BPRS items clustered closer to negative and disorganized aspect of the SIPS. According to the model proposed by Van Os and colleagues these findings could imply that synergism between CBCL and BPRS is related to the fact that two instruments aid in prediction of two independent “affective” and “negative/disorganized” clinical pathways of vulnerability to psychosis [62].

### Limitations

A significant limitation of the current manuscript is that several methodological differences across the two samples hinder the ability to directly compare results of network analysis across 22q11DS and non-syndromic clinical high-risk individuals. Indeed, different clinical instruments, different length of longitudinal follow, different therapeutic strategies and different mean age across the two samples could all contribute to the observed difference in network structure. In this perspective the interest of using independent cohorts was mostly to evaluate the potentialities of our methodological approach in a population that was less genetically and clinically homogenous that 22q11DS, more so than to directly compare candidate clinical pathways across samples. Finally, a significant limitation is that we did not explicitly test for the causal nature of the longitudinal interactions between symptoms. Hence while the structure of such longitudinal correlations remains interesting from the clinical perspective of prognosis, conclusions regarding the existence of causal disease pathways between symptoms remain speculative.

### Future Directions

From a methodological perspective this study should be considered as a proof of concept of the potentialities of graph signal processing techniques to the study of psychopathology. Indeed, both for network dimensionality reduction and graph diffusion we employed the most basic approaches available. It is highly likely that more sophisticated approaches to graph diffusion could improve prediction accuracy and should be the focus of future work.

Moreover, for both populations we reconstructed a single symptom network in the entire sample. While the approach is justifiable in 22q11DS, overall weaker correlations between symptoms observed in the NEURAPRO sample, suggest that additional factors could influence heterogeneous network structure in subgroups of individuals. Interestingly numerous potential hypothesis can be empirically tested in a network framework, including for instance differences in longitudinal network structure with age or across sexes. Finally, while our analysis was limited to clinical scores a network approach is potentially extremely flexible for integrating data originating from different modalities, including for instance neuroimaging or genetics. Embedding a candidate biomarker in the context of longitudinal symptom network could offer an intuitive characterization of clinical variables that are affected. Moreover, the graph diffusion approach could allow to explicitly test the additive value of candidate biomarkers in terms of predictive clinical evolution. Indeed the benchmark against which any future biomarker should be tested is that of improving prediction achieved from gold standard clinical characterization instead of testing prediction performance independently from clinical scores [63]. Crucially, providing additive predictive values implies capturing processes that are not accessible to clinical evaluation more so that describing cross-sectional biomarkers that are strongly correlate with clinical scores, which has been the focus of most current genetic and neuroimaging research [64]. A pragmatic approach could be to investigate whether underlying neurobiological mechanisms are associated with differences in the structure of longitudinal symptoms network and hence improve accuracy of graph-diffusion based prediction.

## Data Availability

Data referred to in the manuscript is available upon reasonable request to the corresponding author.

